# A model of COVID-19 propagation based on a gamma subordinated negative binomial branching process

**DOI:** 10.1101/2020.07.08.20149039

**Authors:** Jérôme Levesque, David W. Maybury, R. H. A. David Shaw

## Abstract

We build a parsimonious Crump-Mode-Jagers continuous time branching process of COVID-19 propagation based on a negative binomial process subordinated by a gamma subordinator. By focusing on the stochastic nature of the process in small populations, our model provides decision making insight into mitigation strategies as an outbreak begins. Our model accommodates contact tracing and isolation, allowing for comparisons between different types of intervention. We emphasize a physical interpretation of the disease propagation throughout which affords analytical results for comparison to simulations. Our model provides a basis for decision makers to understand the likely trade-offs and consequences between alternative outbreak mitigation strategies particularly in office environments and confined work-spaces. Combining the asymptotic limit of our model with Bayesian hierarchical techniques, we provide US county level inferences for the reproduction number from cumulative case count data over July and August of this year.

## 1. Introduction

As of June 20, 2020, there have been more than 8 million confirmed global cases of COVID-19, a respiratory illness caused by the severe acute respiratory syndrome coronavirus 2 (SARS-CoV-2). Early indications suggest a case/infection fatality rate of between 0.5% to 2% [1, 2, 3, 4] with poor prognosis strongly dependent on comorbidity factors such as advanced age, diabetes, and other poor health conditions [5]. The Centers for Disease Control and Prevention in the United States gives an overall current symptomatic case fatality ratio of 0.4% [6] while studies involving seroprevalence indicate a median infection fatality rate of 0.25% [7]. Canada has seen over 100,000 cases and the entire world has engaged in costly outbreak mitigation strategies to prevent excess deaths.

Governments around the world have focused on controlling COVID-19 outbreaks primarily by reducing direct human-to-human contact through varying degrees of society-wide lock-downs and strong social distancing measures. By limiting the opportunity for infectious contacts, the hope is that the infection rate will remain low enough to prevent medical support systems from becoming overwhelmed while also reducing the effective reproduction number of the disease. Evidence suggests that government policies are having a positive effect [8], but some strategies may also become prohibitively expensive in the not too distant future. An alternative outbreak controlling strategy to lock-downs is contact tracing with isolation. In this strategy, health authorities trace the human-to-human contacts of an infected person and isolate those contacts who are at risk of having become infected. If the probability of isolating potentially infected contacts is high and the time to isolation is sufficiently short, contact tracing with isolation may offer better cost benefit performance relative to lock-downs in keeping society safe [9].

Modelling the spread of infectious diseases falls into two broad classes [10]: deterministic modelling, which captures the thermodynamic limit and large scale behaviour of the underlying epidemiological phenomenon, and stochastic modelling, which describes the microscopic statistical nature of the generative process. Traditional compartmental models (e.g., SIRD), usually expressed as a set of coupled ordinary differential equations, fall into the first class while branching processes, in which each infected individual randomly generates “offspring”, belong to the second class. In this paper, we focus on a stochastic formulation of COVID-19 following Hellewell et al. [11].

In the early stages of an epidemic, in a well-mixed and homogeneous population with an infected population that is small relative to the susceptible population, branching models well approximate the dynamics of propagation [12]. A formal mathematical treatment of this statement can be found in [13], and [14], with extensions to explosive growth models found in [15]. Branching model approximations for epidemic application appear at least as early as the mid-1950s [16, 17], and continue to find use in public health domains (see for example, [18, 19, 20]). Branching processes have also found application in modelling past epidemics such as Ebola Virus Disease outbreaks in West Africa [21, 22]. For an introduction to branching processes, see [23], chapter 11, and for a wider introduction to the application of branching processes in other biological fields see [24].

In the current COVID-19 pandemic, branching models have found use in estimating the probability that an imported case into a susceptible population will lead to sustained transmissions [25], and in developing the efficacy of contact tracing strategies [11]. Given the high variance of the observed case count data with COVID-19, especially during the start of a local outbreak, branching process provide and excellent framework for capturing the stochastic nature of super-spreading events [26]. For understanding the challenges in predicting regional trajectories of COVID-19, and for generating relevant policy insights, [27] emphasize the importance of parsimonious models. The authors also stress the usefulness of branching processes, including self-exciting point processes, when analyzing individual COVID-19 count data with parsimonious constructions.

In [11], the authors develop a branching process to model propagation in the presence of contact tracing with isolation. The model uses a negative binomial distribution to generate secondary cases produced by an infected individual with new infections assigned a time of infection through draws from a serial interval distribution. By truncating the serial interval distribution through isolation events, the authors show that in most of their scenarios contact tracing and case isolation is enough to control a new outbreak of COVID-19 within 3 months.

While the construction in [11] provides a rich base for numerical simulations, to gain further insight, we extend the model to a fully continuous time setting which provides us with a complete generative model, including expressions for the generating and characteristic functions. Furthermore, each part of our model has a direct physical interpretation of the underlying disease propagation mechanism. The model balances fidelity and parsimony so that the model can 1) be calibrated to data relatively easily, 2) provide semi-analytic tractability that allows for trade-off analysis between different mitigation strategies 3) generate realistic simulated sample paths for comparing interventions. Our code is available as R packages^1 2^.

## 2. The model

In this paper, we build a Crump-Mode-Jagers (CMJ) branching process model through a subordinated Lévy process. CMJ constructions contain the triple of random processes (*λ*_*x*_, *ξ*_*x*_(·), *χ*_*x*_(·)) defined by,

- *λ*_*x*_ is a random variable that denotes an infected person’s communicable period;
- *ξ*(*t*) = #*{k* : *σ*(*ω, k*) ≤ *t}* counts the number of infected people over event space Ω(*ω*) in time *t. ξ*_*x*_(*t* − *σ*_*x*_) denotes the random number of infected people created by an infected person at every moment of her communicable period over the interval [*σ*_*x*_, *t*); *ξ*(*t* − *σ*_*x*_) = 0 if *t* − *σ*_*x*_ *<* 0; and
- *χ*_*x*_(*t* − *σ*_*x*_) is a random characteristic of the infected person within the interval [*σ*_*x*_, *t*); *χ*(*t* − *σ*_*x*_) = 0 if *t* − *σ*_*x*_ *<* 0. (E.g., *χ*(*t*) = 𝕀 {*t* ∈ [0, *λ*_*x*_)} is the number of infectious existing at moment *t*).

Our model begins by generating infections from an infected individual through a compound Poisson process where the event times represent transmission events (*σ*_*x*_). We assume that an individual is infectious from the moment she becomes infected. The number of new infections at each transmission event is a draw from the logarithmic distribution [28] (*ξ*(*t*)) and consequently, the resulting generative process is the negative binomial process (see, for example, Quenouille [29]). The stochastic counting processes remains “on” during the communicable period and then shuts “off” at the end—that is, the communicable period is the random lifetime (*λ*_*x*_) in the CMJ language. We model the communicable period as a gamma distributed random variable, Γ(*a, b*), with mean 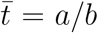. By subordinating our resulting negative binomial process with a gamma process for the communicable period, we arrive at our model of COVID-19 propagation—a gamma negative binomial branching process (GNBBP). (For details on subordinated Lévy processes, see [30].)

The model we present assumes a well-mixed population in that each child in the branching processes has the same statistical properties as the parent. While our model includes the property of super-spreader events, generated by the fat tail of the counting process, it does not include heterogeneous mixing scenarios whereby susceptible contacts for an infected individual diminishes each time a new person becomes infected. Furthermore, the branching model assumes an infinite reservoir of susceptible people for the virus to spread (the process branches without regard to population constraints). This approximation holds provided that the susceptible population is not near exhaustion. We focus on the early days of an outbreak with infected population sizes small compared to the total susceptible population over time scales not hierarchically larger than the communication window. It is this limit that is most relevant to decision makers for setting workplace policies.

### 2.1. Construction details

We model the propagation of COVID-19 by assuming that people become infectious immediately after contracting the virus and that they can infect others throughout the duration of their communicable period. We assume the population is homogeneous and that each new infected individual has the same statistical properties as previously infected people. Specifically, we assume that an infected person infects *Q*(*t*) other people during the given time interval [0, *t*] according to a compound Poisson process,

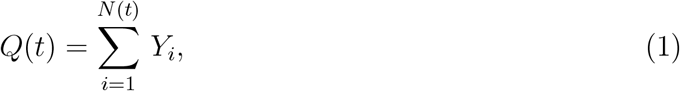

where the number of infectious events, *N* (*t*), follow a Poisson counting process with arrival rate *λ*, and *Y*_*i*_, the number infected at each event, follows the logarithmic distribution,

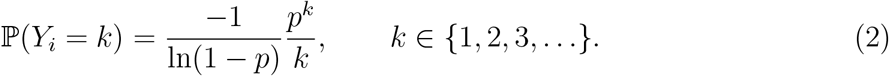

The characteristic function for *Q*(*t*) reads,

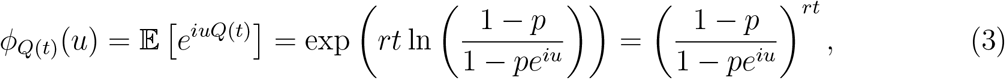

with *λ* = −*r* ln(1 − *p*); thus *Q*(*t*) follows a negative binomial process,

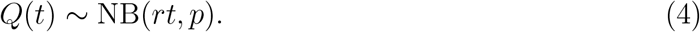

In this process, during a communicable period, *t*, an infected individual infects *Q*(*t*) people based on a draw from the negative binomial with mean *rtp/*(1 − *p*). The infection events occur continuously in time according to the Poisson arrivals. However, the communicable period, *t*, is in actuality a random variable, *T*, which we model as a gamma process^3^ with density,

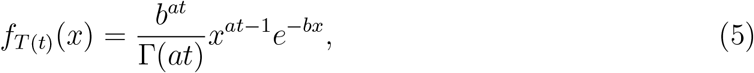

which has a mean of 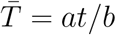. By promoting the communicable period to a random variable, the negative binomial process changes into a Lévy process with characteristic function,

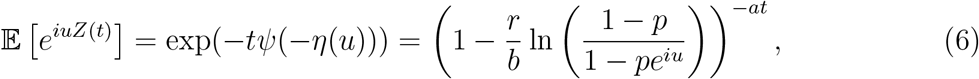

where *η*(*u*), the Lévy symbol, and *ψ*(*s*), the Laplace exponent, are respectively given by,

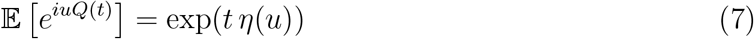

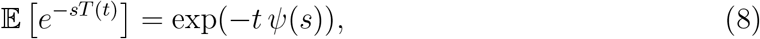

with,

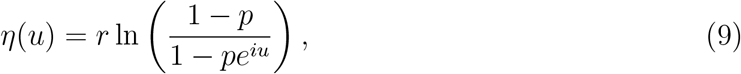

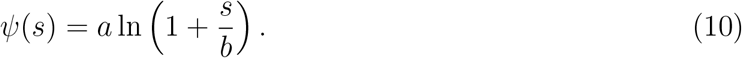

*Z*(*t*) is the random number of people infected by a single infected individual over her random communicable period. Without loss of generality, we absorb *t* into *a* (or alternatively set *t* = 1, representing a single lifetime) giving a mean communicable period 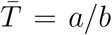. The gamma process smears out the end of the communicable period.

We see that *R*_0_ = 𝔼 [*Z*(1)] = *arp/*(*b*(1 − *p*)), and thus our process has the same mean as the negative binomial process with a fixed stopping time of *t* = *a/b*. In fact, since *λ* = −*r* ln(1 − *p*) we have the simple relationship,

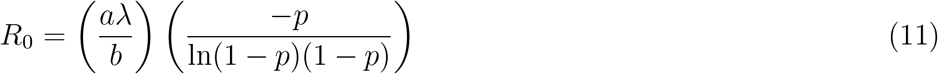

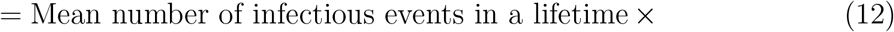

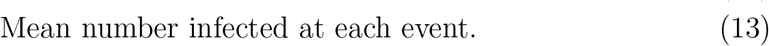

The variance of the our counting process is over-dispersed relative to the the negative binomial,

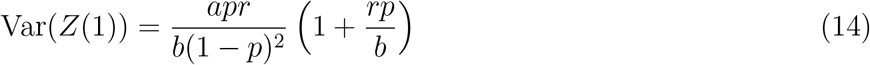

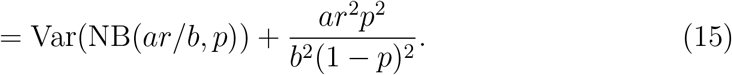

The model has four parameters:

- *p* sets the number of infected people per infectious interaction. The mean number of infected people per infectious event is, 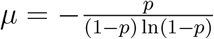.
- *λ* = −*r* ln(1 − *p*) gives the arrival rate of infectious events.
- *a, b* together set the mean communicable period, 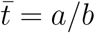, and determine the variance along with the skewness and kurtosis of the gamma distribution, Γ(*a, b*). In the limit *b* → 0 with *a/b* finite, the gamma distribution becomes a delta function at the mean time and we recover the negative binomial process evaluated at *t* = *a/b*.

The characteristic function eq.(6) of the stopped stochastic process allows us to explore the model’s analytical properties, which can help decision makers better understand trade-offs in small environments.

### 2.2. Renewal equations and Malthusian parameters

Given a random characteristic *χ*(*t*), such as the number of infectious individuals at time *t*, (e.g., *χ*(*t*) = 𝕀 (*t* ∈ [0, *λ*_*x*_)) where *λ*_*x*_ is the random communicable period) the expectation of the process follows,

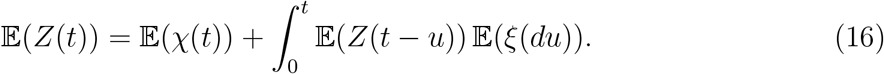

Defining the Malthusian parameter, *α >* 0, if it exists, by,

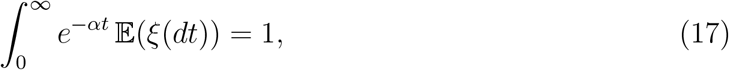

we can change eq.(16) into a renewal equation,

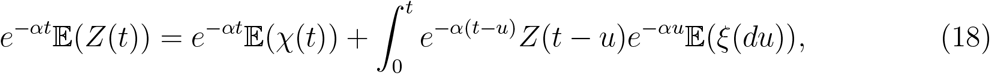

which has the solution,

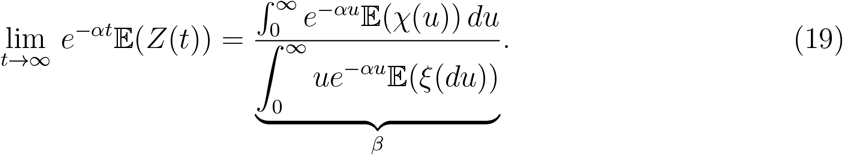

Thus the asymptotic behaviour of the solution is governed by the pair parameters *α*, and *β*.

Recall that *ξ*(*t*) = # {*k* : *σ*(*ω, k*) ≤ *t*} counts the number of infections during the observation window [0, *t*] over event space Ω(*ω*). In our model we have,

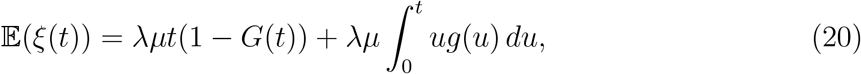

where *λ* and *µ* are respectively the Poisson arrival rate and the mean of logarithmic distribution, and where,

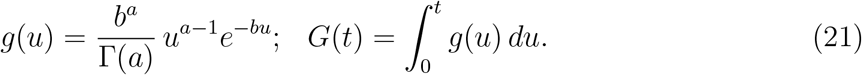

Therefore,

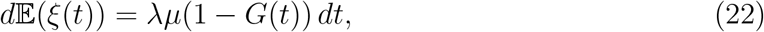

which leads to the expected result for the mean of direct infections per individual,

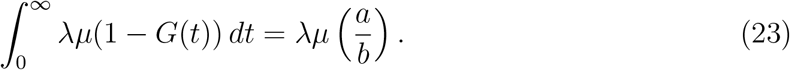

Using eq.(22) and eq.(17) we find that,

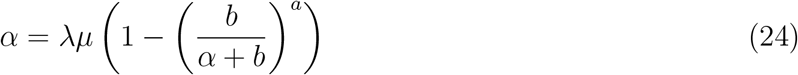

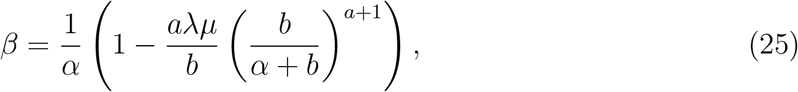

which we can solve for the Malthusian parameter, *α*, by Newton-Raphson. The asymptotic solution to eq.(16) given that the Malthusian parameter exists is,

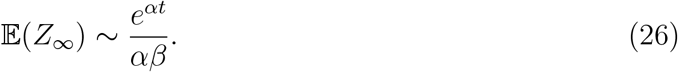

If *R*_0_ *<* 1 the branching process will not experience asymptotic exponential growth, instead we can solve eq.(16) for its long term limit,

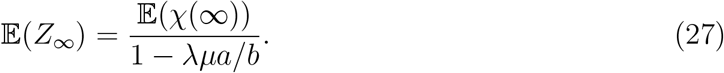

### 2.3. Extinction probabilities and component sizes

In the CMJ framework, we have the generating function,

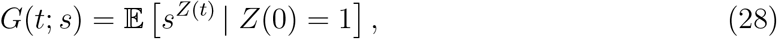

with the number of infected by time *t* composed of infections at time *σ*_*i*_,

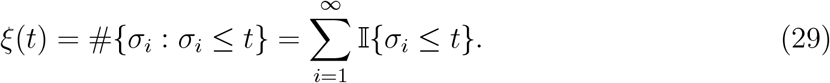

The probability of extinction reads,

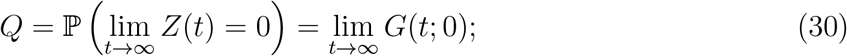

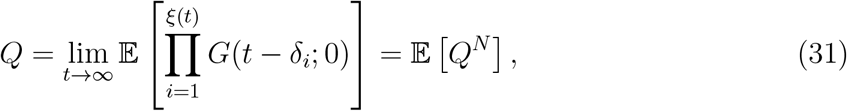

and for our model, we arrive at the transcendental relationship,

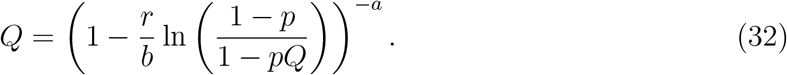

Again, we can apply Newton-Raphson and solve for the extinction probability *Q*.

In addition to the extinction probability for our branching process, we can estimate the average number of total infected people at extinction if extinction occurs by considering the theory random graphs. The branching process is a directed bipartite graph (it is a tree) and given the generating function for the process, we know the distribution of the outgoing edges from a randomly chosen vertex. In [31], the authors extend Erdos-Renyi constructions of random graphs to graphs with arbitrary vertex degree. They compute the mean component size for graphs, including graphs excluding the giant component, if it exists.

The total number infected corresponds to the random characteristic 𝔼(*χ*(*t*)) = 1 and thus eq.(27) has the non-Malthusian growth solution,

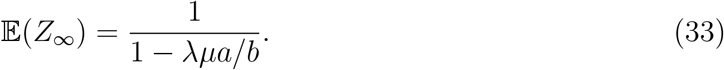

In [31], the authors consider two generating functions,

- *G*_0_(*s*): the generating function for the probability distribution of the vertex’s degree; and
- 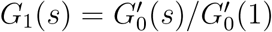: the generating function for the probability distribution of the outgoing edges from a randomly chosen vertex.

Eq.(6) with *e*^*iu*^ → *s* is *G*_1_(*s*) in the notation of [31] and for our purposes we do not need an explicit formula for *G*_0_(*s*). The average component size of the graph, in the absence of a giant component, is [31]

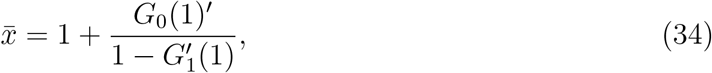

which matches the renewal equation solution eq.(33) if 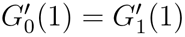—that is, the generating two functions intersect tangentially at *s* = 1.

At 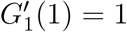 a phase transition occurs and the giant component emerges. The fraction of the graph occupied by the giant component is,

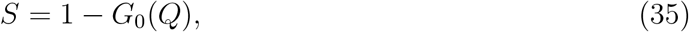

where *Q* is the extinction probability for the distribution of outgoing edges, *Q* = *G*_1_(*Q*). Since the fraction of the graph that does not belong to the giant component is composed precisely of those graphs which have gone extinct in our process, we impose *G*_0_(*Q*) = *Q*. Thus, we demand that the two generating functions intersect on the 45° line at *Q <* 1 when *α >* 0. The average component size in this case becomes [31],

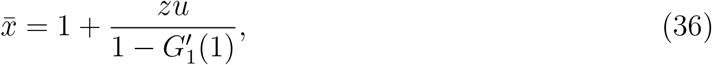

where,

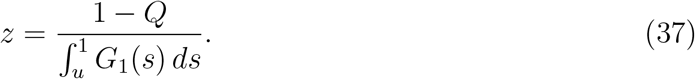

As *Q* → 1 we see that *G*_1_(*Q*) and *G*_0_(*Q*) increasingly intersect tangentially, finally becoming tangent at *Q* = 1, which is consistent with our observation in eq.(34). We take 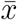 to be the average size of the total infected population at extinction, if extinction occurs.

## 3. Contact tracing and propagation interruption

The process in eq.(6) represents the spread of the disease from an infected individual without any mitigation strategies. Imagine that we can trace, contact, and isolate infected individuals with a success probability *q* and with an mean time to isolation of 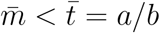 after the infectious event. We assume that once isolated, there is no chance for the infected individual to spread the disease any further. We again imagine that the isolation time is gamma distributed but with parameters (*a*′, *b*′) leading to the isolation process, 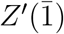,

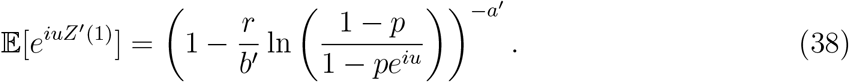

Notice that the branching process for a successful isolation event has the same form as the original process with a mean time of the random communicable period of 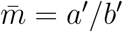. Thus, the trace-contact-isolate branching process becomes,

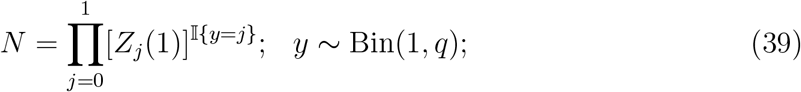

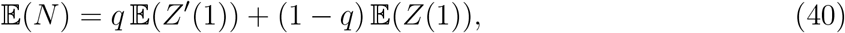

where *N* is the number of infections produced by an infected person during her communicable period, and *q* is the probability of a successful isolation event. Instead of arbitrarily cutting the communicable period’s density function based on an isolation event as prescribed in [11], our model maintains the same form of the generating function throughout by shifting the mean and variance of the communicable period’s gamma process. In a contact-trace-isolate policy, the expected number of infections per infected individual becomes,

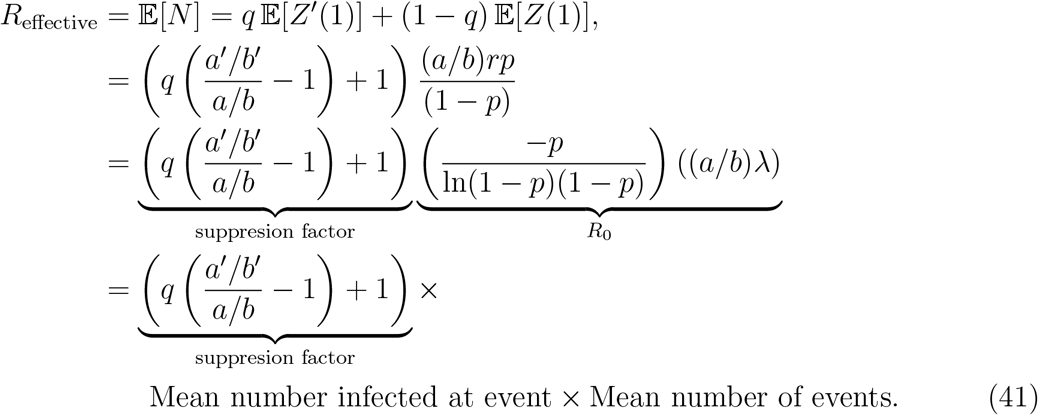

Eq.(41) provides intuition for comparing competing courses of action by affording trade-off analyses. In a lock-down, health authorities control the spread of the disease by lowering the human-to-human interaction rate *λ*. If *λ* can be made sufficiently small, *R*_0_ will drop below unity and the outbreak will come under control. The first term in eq.(41) represents a suppression factor, which by construction is less than unity, and results from an isolation policy with success probability *q*. Alternatively, that same reduction in *R*_effective_ can also be achieved by a lock-down scenario if the infection event rate, *λ*, is reduced^4^ by the same suppression factor. Thus, we see an equivalence in generating *R*_effective_ from the two different mitigation strategies, each of which may come at different economic costs.

To make the observation concrete, suppose *λ* = .20 implying an average of 0.20 infectious events per day, *p* = 0.5 implying an average of 1.44 infections per infectious event, and a mean communicable period of *a/b* = 5.5 days. The parameters imply *R*_0_ = 1.59. Figure 1 shows iso-contours of fixed suppression factor in the *a*′*/b*′ − *q* plane. We can now see the trade-off between a lock-down policy with a fixed suppression factor and a contact tracing with isolation policy which generates the suppression factor from successful contact tracing events. The economic costs of generating the same value the suppression factor among the two strategies (lock-down vs contact tracing with isolation), with its corresponding reduction in *R*_effective_, will in general not be the same. From the figure we see in this example that a contact tracing with isolation policy with an isolation probability of 0.75 and a mean isolation time of 4 days is equivalent to reducing the rate of human-to-human infection events by a factor of approximately 1.25. A lock-down that reduces human interactions by a factor of 1.25 will almost certainly cost much more than the corresponding contact tracing with isolation strategy [9].

**Figure 1:**
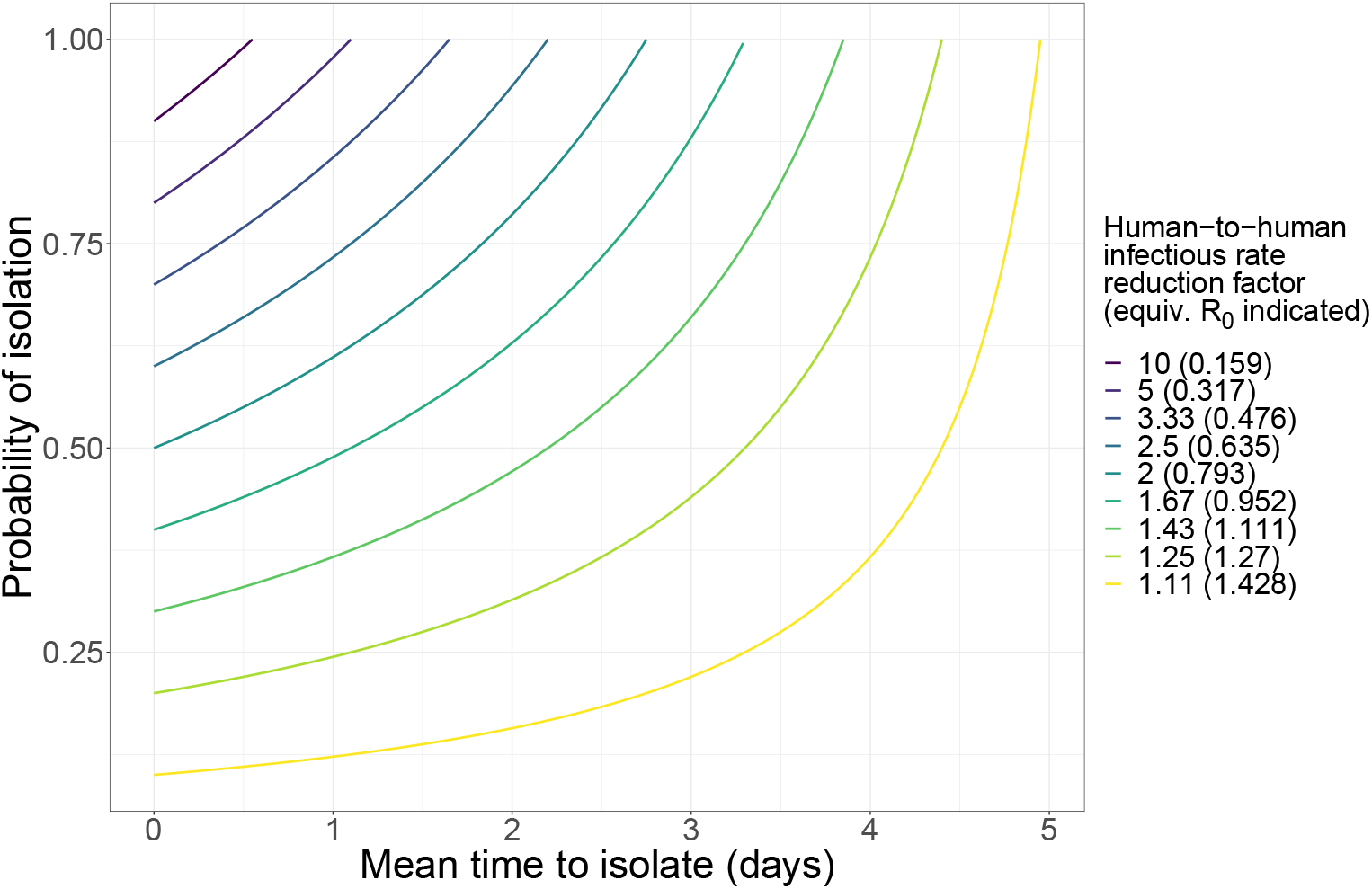
Iso-contours in the *a*′*/b*′ − *q* plane with equivalent lock-down factor *f*.

## 4. A scenario planing exercise: Policy input for return to work

One area of application for our model is helping decision makers understand counterfactual outcomes in a return-to-work policy exercises. In setting policies, decision makers must weigh the operational needs of their business against the possibility of an outbreak in the work environment. In addition to the analytical results that our model provides, simulation can further help ring-fence difficult decisions.

Our model requires four parameters, the arrival rate of infectious interactions, the average number infected at each event, and two parameters which govern the communicable period’s density function. We use the open literature [32] as a guide to fix the communicable period; we fix *a* = 4.7 days with *b* = 0.85; these parameter choices give a mean communicable period of 5.5 days with 97.5% of the communicable period ending in 11.5 days. In figure 2, we show the density function arising from our parameter settings. The decision maker has control over the remaining two parameters. By limiting meeting sizes, restricting the number of employee interactions, and by mandating the use of personal protective equipment, the decision maker can set the variables controlling the arrival rate of infectious events and the number infected at each event. We treat the population as homogeneous and well-mixed, holding fixed the arrival rate for infections and the number infected per event over time. In a small setting, in reality, we expect that as people become infected the social network will change, even in the limit of an unmitigated outbreak. We eventually expect heterogeneity to occur and a breakdown of the well-mixed assumption. Those changes will have an effect on the basic parameters of our branching model as the population becomes infected, but exactly how the network changes is a complicated phenomena. Feedback can move the arrival rate and the number infected at each event in competing directions. By ignoring any time dependence in the basic parameters, our model provides a baseline understanding on how COVID-19 propagates in a small well-mixed populations and over time scales not hierarchically larger than the communicable period.

**Figure 2:**
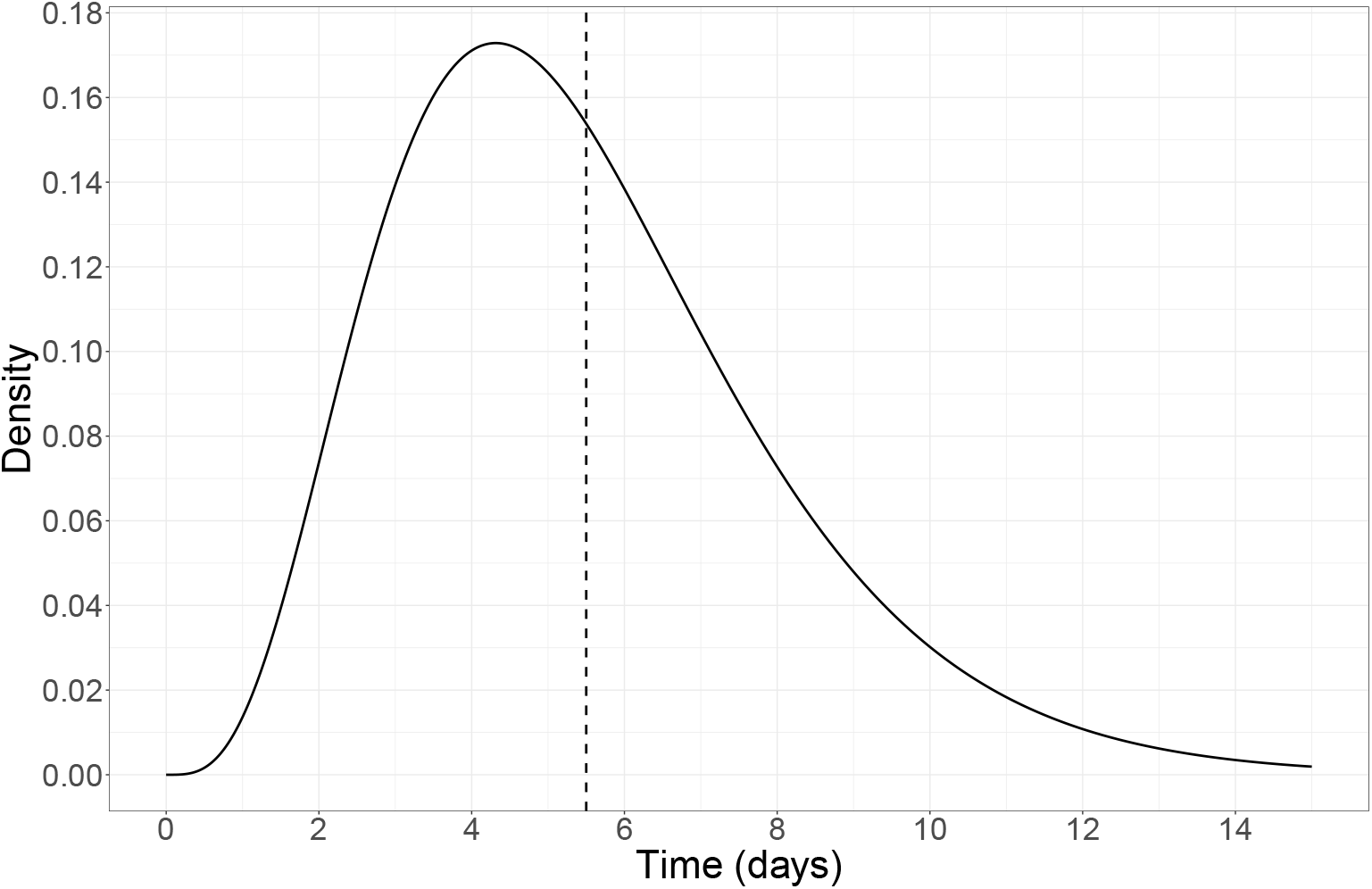
The communicable period: *a* = 4.7, *b* = 0.85. The mean of 5.5 days is indicated by the read vertical line.

Imagine a scenario in which a decision maker has an office space with 100 employees and she must decide on mitigation strategies. Suppose a baseline scenario with *λ* = 0.2 and *p* = 0.5 (corresponding to an average of 1.44 infected per event). Given the properties of the communicable period, this scenario corresponds to *R*_0_ = 1.59, which implies that if an infected person arrives in the population, in expectation, the branching process will lead to exponential growth in infections. In figures 3a and 3b we display the solution to the renewal equation with this parameter choice for the expected number of infected people and the expected size of the active infectious population respectively. Let us suppose that the decision maker can change the model parameters *λ* and *p* through policy considerations, creating two possible alternative scenarios, each coming at different financial costs. In scenario 1, the manager can control meeting sizes but not the interaction rate of her employees. As a result, she reduces the logarithmic distribution parameter by a factor of 2 which translates to a 20% reduction in the mean of the logarithmic distribution. Scenario 2 is the reverse of scenario 1—the manager can reduce the interaction rate of the employees, but not the meeting sizes. We imagine that in scenario 2 the manager cuts the employee interaction rate in half. Our model allows the decision maker to investigate trade-offs between these scenarios all starting from one undetected infected individual in the workplace. We summarize model outputs between two scenarios in table 1. We see that in scenario 2, the manager’s policy reduces *R*_effective_ below unity indicating that if an infected employee started a chain of infections, the propagation would extinguish on its own. In this scenario, the mean number of infected people after two weeks is only 3.3. These observations can also help the manager decide on floor occupancy levels, given how far the virus would propagate over two weeks in the limit of a well mixed homogeneous population in a large susceptible background.

**Table 1:** Model properties of the planning scenarios. Each scenario starts with one undetected infected individual.

| description | symbol | scenario properties |  |  |
| --- | --- | --- | --- | --- |
|  |  | baseline | scenario 1 | scenario 2 |
| Poisson arrival rate | $\lambda$ | 0.20 | 0.20 | 0.1 |
| Logarithmic distribution parameter | $p$ | 0.50 | 0.15 | 0.5 |
| communicable period shape parameter | $a$ | 4.7 | 4.7 | 4.7 |
| communicable period rate parameter | $b$ | 0.85 | 0.85 | 0.85 |
| mean number of new infections per infected individual | $R_0$ | 1.59 | 1.28 | 0.80 |
| extinction probability | $Q$ | 0.60 | 0.71 | 1 |
| mean size at extinction | $\bar{x}$ | 3.1 | 4.7 | 4.9 |
| mean number infected after one week | $N_{1w}$ | 6 | 4 | 2.3 |
| mean number infected after two weeks | $N_{2w}$ | 22 | 10 | 3.3 |

**Figure 3:**
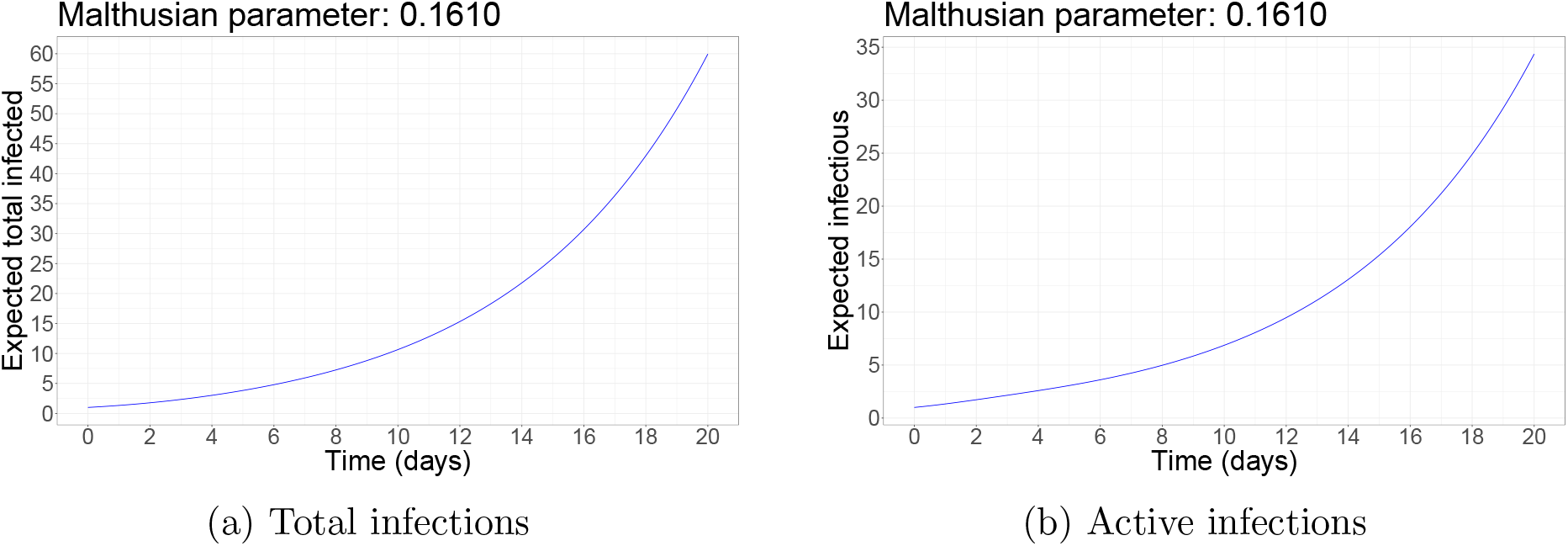
The expected number of total and active infections as function of time in the baseline planning scenario.

In some office environments, we can also imagine a scenario in which management introduces an aggressive testing scheme to isolate infected employees. Suppose our manager faces the baseline scenario in table 1 but instead of manipulating the interaction rate or meeting sizes, the manager implements a test with a 90% chance of a successful isolation that is sharply peaked around a mean of 0.69 days. We approximate the effect of testing with a successful isolation event by setting *a*′ = 1.17, and *b*′ = 1.7 in eq.(41) and we show the comparison to the baseline density function established from [32] in figure 4. Using eq.(41) we see that *R*_effective_ = 0.21 *×* 1.59 = 0.33, and thus this isolation strategy turns an exponentially growing configuration into a process that will go extinct almost surely. Figure 5 shows 1,000 sample paths of the isolation process over 30 days. Most paths go extinct within ten days and the average total number of infected is 3.8 people.

**Figure 4:**
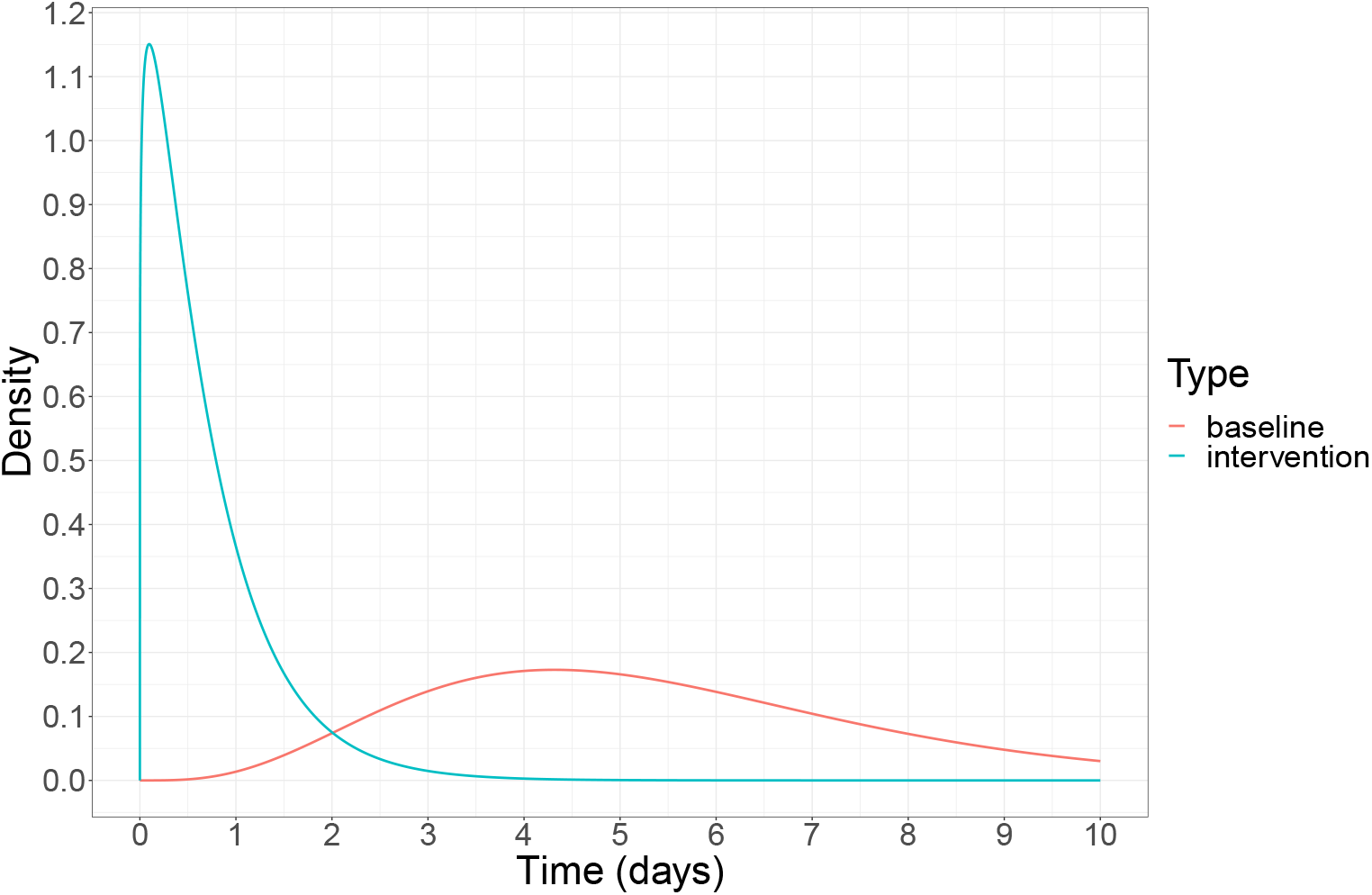
The communicable period density function with a successful isolation event (*a* = 1.17, *b* = 1.7) compared to the baseline scenario.

**Figure 5:**
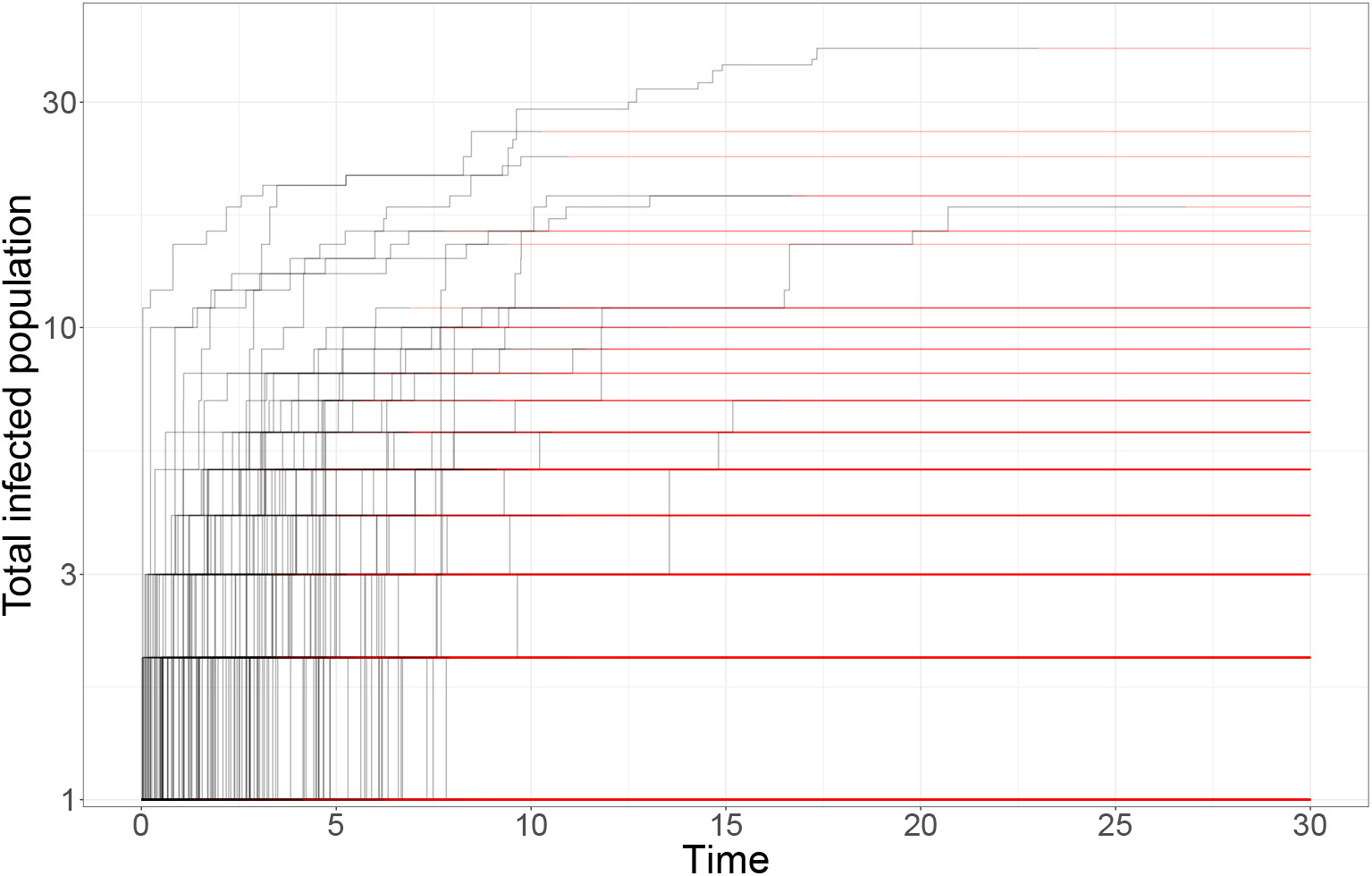
A contract tracing with isolation strategy: 1,000 simulations over 30 days. The probability of successful isolation is *q* = 0.90, with gamma distribution parameters *a* = 1.17, and *b* = 1.7. Red lines indicated extinct paths from the moment of extinction. All paths eventually go extinct as the result of the intervention.

## 5. A note on parameter inference and an example with US county data

This paper describes a gamma negative binomial branching process (GNBBP) on the number of new infections generated by an infected individual. Given a set of observed 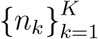 infection counts for *K* individuals, a complete Bayesian analysis of the model is possible, in which all model parameters are identifiable, using for example, the infrastructure provided in [33]. Under this scheme, all four parameters (*r, p, a, b*) can be resolved, allowing for a full posterior predictive analysis.

Define a complete history of an outbreak as a set of *N* observations taking the form of a 6-tuple:

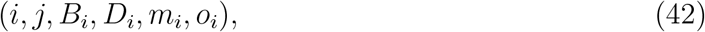

where

*i* index of individual

*j* index of parent

*B*_*i*_ time of birth

*D*_*i*_ time of death

*m*_*i*_ number of offspring birth events

*o*_*i*_ number of offspring.

With the following summary statistics

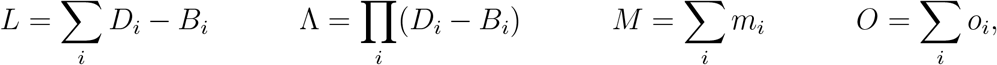

we can build a Gibbs sampler over the GNBBP parameters as follows:

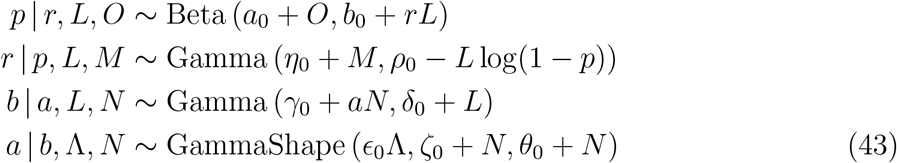

where *a*_0_, *b*_0_, *η*_0_, *ρ*_0_, *γ*_0_, *ζ*_0_, *ϵ*_0_, *θ*_0_ are hyper-parameters.

Unfortunately, under real world conditions, we are rarely fortunate enough to have such complete and pristine temporal data. In practice, it will almost never be possible obtain such complete histories, but the Gibbs sampler construction shows that the model is identifiable from complete data. Readily available COVID-19 data almost always takes the form of cumulative case count data by geographic region, but if public health officials can collect data in the form of eq.(42) during a local outbreak, Gibbs sampling will yield posteriors for all model parameters. (An efficient contact tracing strategy for an outbreak in correctional facilities, or isolated northern Canadian and Indigenous communities might reveal reliable temporal patterns. If such strategies could be implemented, the data would allow Bayesian inference of the entire model.) The underlying propagation mechanism of the GNBBP affords additional interpretability to the model, which, in turn, facilitates incorporation of other prior information. For example, knowing that it is unlikely that multiple thousands of individuals could be infected in a single interaction allows us to set a prior with more mass on values of *p* closer to 0; moreover, direct experimental measurement of this parameter might be possible in a laboratory setting or augmented by fine grained clinical data. Similar considerations apply to the rate of infectious events, *λ*. The parameters which govern the communicable period, (*a, b*), can be inferred from clinical observations. Likewise, information on probable ranges of *R*_0_ from other comparable infections could also be leveraged to provide a joint constraint on *r, p, a*, and *b*.

Even with limited data we can still estimate parts of the model. In particular, we can estimate the Malthusian parameter of eq.(17) from cumulative count data that exhibits exponential growth by applying the asymptotic solution, eq.(26). Since the Malthusian parameter depends on the product of the infection arrival arrival rate and the average number infected per event, *λµ*, an estimate of the Malthusian parameter yields an estimate of *R*_effective_ through the parameters *a* and *b* of the communicable period’s gamma distribution,

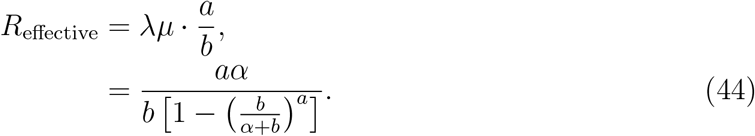

Based on the clinical literature [32], we take *a* = 4.66 and *b* = 0.85 giving a mean communicable period of approximately 5.5 days with a 97.5% of 11.5 days.

The New York Times provides a COVID-19 case count dataset for the United States resolved on the county level [34]. A team at the New York Times curates the data from multiple sources and ensures data accuracy. Using the New York Times data, we estimate the Malthusian parameter for US counties which exhibit exponential growth over the period July 1, 2020 to August 20, 2020. We use a hierarchical Bayesian construction with a county level random effect,

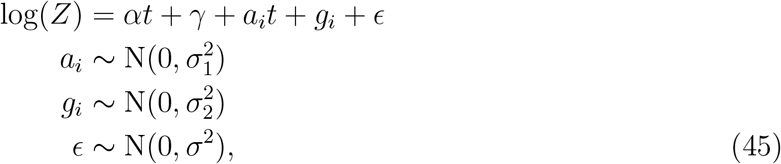

where *i* is the county label; the variance parameters use half-Cauchy priors and the fixed and random effects use normal priors. Table 2 summarizes the variables of the linear mixed effects inference model. We estimate the model and generate posterior distributions for all parameters using JAGS [35]. Our code is publicly available in an R package. The posterior means of the Malthusian parameter for each county gives *R*_effective_ over the time interval through eq.(44). In figure 6, we display the US county results for *R*_effective_. Over the mid-summer, we see that the geographical distribution of *R*_effective_ across the US singles out the Midwestern states and Hawaii as hot-spots while Arizona sees no county with exponential growth.

**Table 2:** Linear mixed effects model parameters for calibrating the GNBBP model to US county data.

| Variable | Symbol |
| --- | --- |
| Fixed effect slope (Malthusian parameter) | $\alpha$ |
| Fixed effect intercept | $\gamma$ |
| County level slope random effect | $a_i$ |
| County level intercept random effect | $g_i$ |
| Slope random effect variance | $\sigma_1^2$ |
| Intercept random effect variance | $\sigma_2^2$ |
| Model variance | $\sigma^2$ |

**Figure 6:**
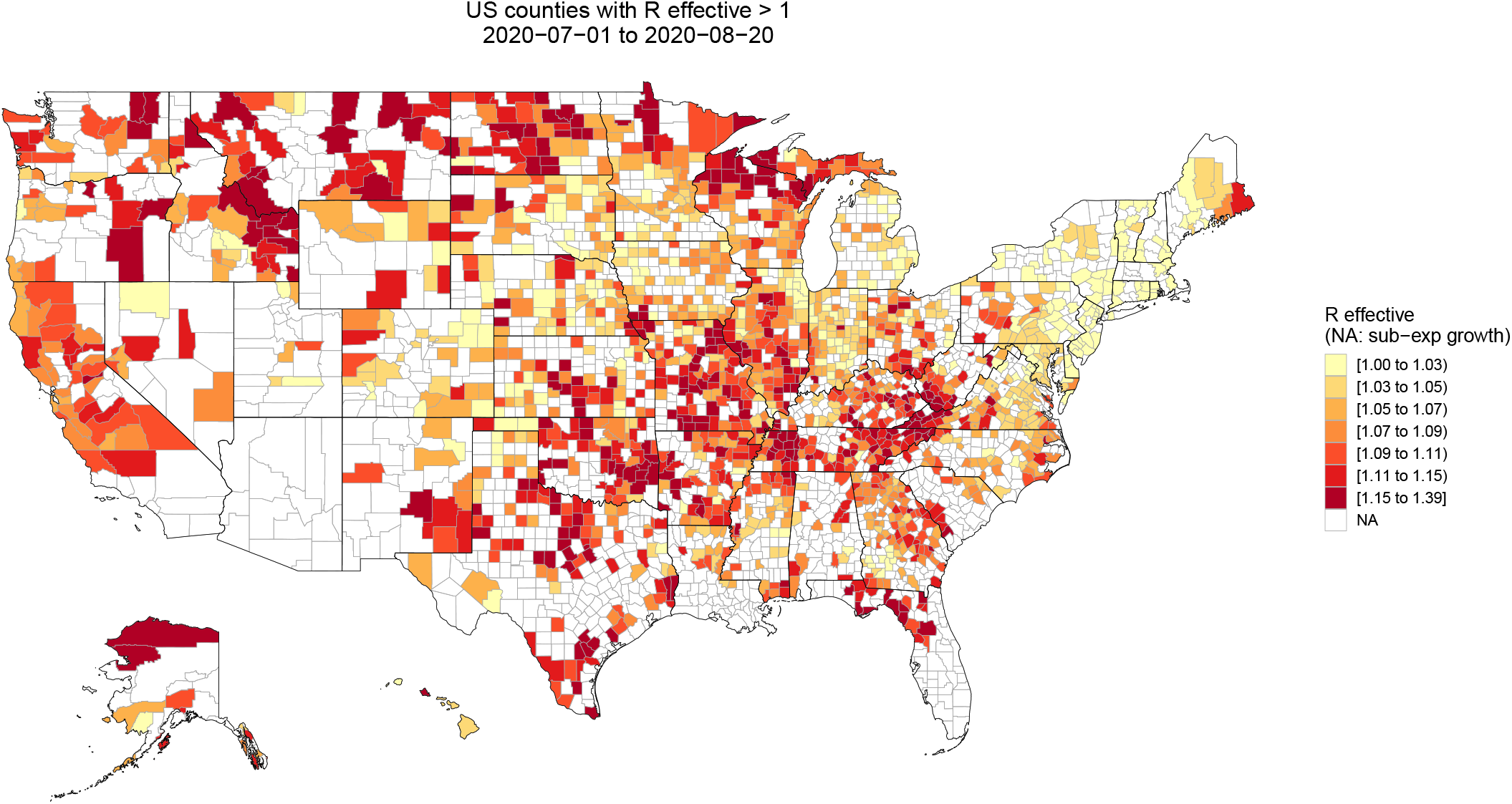
Summer 2020 geographical distribution of *R*_effective_ across the United States: 2020-07-01 to 2020-08-20.

## 6. Discussion

In small settings with localized outbreaks, a branching model offers a stochastic view of the propagation. To be useful in a decision making setting, the branching model must be parsimonious yet contain appropriate features which match clinical observations and bounds on key parameter such as *R*_effective_.

Our model contains physically motivated mechanisms that link to macroscopic observables. For instance, our model generates the negative binomial count process by coupling Poisson infectious event arrivals with the logarithmic distribution for the number infected at each event. We extend the model of [11] by including the serial interval distribution within a complete generative continuous time stochastic branching process. Furthermore, our model allows for an exploration of trade-offs between mitigation strategies at the microscopic level, especially in light of the model’s analytical tractability. Because our model includes the generating function of the underlying branching process, it easy to build a continuous time simulation engine, model the effect of intervention strategies, and estimate model parameters through Bayesian hierarchical methods.

We see application of this model in workspace settings to help management set risk tolerances. Throughout, we assume that the condition of a well-mixed homogeneous population holds. We also assume that the infected population remains small relative to the entire workforce under consideration. The scenario of greatest concern is the arrival of a single asymptomatic person in the workplace who begins a chain of employee-to-employee infections which is only discovered by management through the appearance of the first symptomatic case days or weeks after the initial transmission. This branching model gives a counterfactual sense of how far the virus could spread in the time frame until first discovery, which in turn can help set new polices around employee interactions, meeting sizes, floor occupancy, and physical distancing. The condition under which the branching approximations fail to hold, in that an order one fraction of the population has become infected by first detection, puts the outbreak in a new regime. Those situations would consist of very small initial populations, on the order of the basic reproduction number, or on time frames much longer than the communicable window. However, a large unmitigated outbreak in a small workplace presents entirely new problems, requiring aggressive triage, and will be immediately apparent to management.

## Data Availability

NA

## Footnotes

1 https://github.com/pspc-data-science/branchsim.git

2 https://github.com/pspc-data-science/branchestimate.git

3 We apply a gamma process subordinator to the negative binomial process.

4 We recognize that a lock-down would probably reduce *p* in the logarithmic distribution as well, but we suspect that effect is secondary. We suspect that *p*, which sets the number of people infected during an event, is not nearly as sensitive to a lock-down scenario as compared to the expected number of events during the communicable period.

## References

[1] Robert Verity, Lucy C Okell, Ilaria Dorigatti, Peter Winskill, Charles Whittaker, Natsuko Imai, Gina Cuomo-Dannenburg, Hayley Thompson, Patrick GT Walker, Han Fu, et al. Estimates of the severity of coronavirus disease 2019: a model-based analysis. The Lancet infectious diseases, 2020.

[2] Marc Lipsitch. Estimating case fatality rates of covid-19. The Lancet Infectious Diseases, 2020.

[3] John P. A. Ioannidis, Cathrine Axfors, and Despina G. Contopoulos-Ioannidis. Population-level covid-19 mortality risk for non-elderly individuals overall and for non-elderly individuals without underlying diseases in pandemic epicenters. medRxiv, 2020.

[4] Eran Bendavid, Bianca Mulaney, Neeraj Sood, Soleil Shah, Emilia Ling, Rebecca Bromley-Dulfano, Cara Lai, Zoe Weissberg, Rodrigo Saavedra, James Tedrow, Dona Tversky, Andrew Bogan, Thomas Kupiec, Daniel Eichner, Ribhav Gupta, John Ioannidis, and Jay Bhattacharya. Covid-19 antibody seroprevalence in santa clara county, california. medRxiv, 2020.

[5] Tianbing Wang, Zhe Du, Fengxue Zhu, Zhaolong Cao, Youzhong An, Yan Gao, and Baoguo Jiang. Comorbidities and multi-organ injuries in the treatment of covid-19. The Lancet, 395(10228):e52, 2020.

[6] Covid-19 pandemic planning scenarios. https://www.cdc.gov/coronavirus/2019-ncov/hcp/planning-scenarios.html. Accessed: 2020-06-20.

[7] John Ioannidis. The infection fatality rate of covid-19 inferred from seroprevalence data. medRxiv, 2020.

[8] The Lancet. Sustaining containment of covid-19 in China. Lancet (London, England), 395(10232):1230, 2020.

[9] Daron Acemoglu, Victor Chernozhukov, Ivn Werning, and Michael D Whinston. Optimal targeted lockdowns in a multi-group sir model. Working Paper 27102, National Bureau of Economic Research, May 2020.

[10] Linda JS Allen. A primer on stochastic epidemic models: Formulation, numerical simulation, and analysis. Infectious Disease Modelling, 2(2):128–142,2017.

[11] Joel Hellewell, Sam Abbott, Amy Gimma, Nikos I Bosse, Christopher I Jarvis, Timothy W Russell, James D Munday, Adam J Kucharski, W John Edmunds, Fiona Sun, et al. Feasibility of controlling covid-19 outbreaks by isolation of cases and contacts. The Lancet Global Health, 2020.

[12] Odo Diekmann, Hans Heesterbeek, and Tom Britton. Mathematical tools for understanding infectious disease dynamics, volume 7. Princeton University Press, 2012.

[13] Frank Ball and Peter Donnelly. Strong approximations for epidemic models. Stochastic processes and their applications, 55(1):1–21, 1995.

[14] Peter Jagers et al. Branching processes with biological applications. Wiley, 1975.

[15] Júlia Komjáthy. Explosive crump-mode-jagers branching processes. arXiv preprint 1602.01657, 2016.

[16] David G Kendall. Deterministic and stochastic epidemics in closed populations. In Proc. 3rd Berkeley Symp. Math. Statist. Prob, volume 4, pages 149–165, 1956.

[17] Maurice Stevenson Bartlett. An introduction to stochastic processes: with special reference to methods and applications. CUP Archive, 1978.

[18] Frank Ball, Miguel González, Rodrigo Martínez, and Maroussia Slavtchova-Bojkova. Total progeny of crump-mode-jagers branching processes: An application to vaccination in epidemic modelling. In Branching Processes and Their Applications, pages 257–267. Springer, 2016.

[19] Frank Ball and Laurence Shaw. Inference for emerging epidemics among a community of households. In Branching Processes and Their Applications, pages 269–284. Springer, 2016.

[20] CP Farrington, MN Kanaan, and NJ Gay. Branching process models for surveillance of infectious diseases controlled by mass vaccination. Biostatistics, 4(2):279–295, 2003.

[21] Stefano Merler, Marco Ajelli, Laura Fumanelli, Marcelo FC Gomes, Ana Pastore y Piontti, Luca Rossi, Dennis L Chao, Ira M Longini Jr, M Elizabeth Halloran, and Alessandro Vespignani. Spatiotemporal spread of the 2014 outbreak of ebola virus disease in liberia and the effectiveness of non-pharmaceutical interventions: a computational modelling analysis. The Lancet Infectious Diseases, 15(2):204–211, 2015.

[22] John M Drake, RajReni B Kaul, Laura W Alexander, Suzanne M ORegan, Andrew M Kramer, J Tomlin Pulliam, Matthew J Ferrari, and Andrew W Park. Ebola cases and health system demand in liberia. PLoS Biol, 13(1):e1002056. 2015.

[23] Samuel Karlin. A first course in stochastic processes. Academic press, 2014.

[24] M Kimmel and DE Axelrod. Branching processes in biology., 2002.

[25] Robin N Thompson. Novel coronavirus outbreak in wuhan, china, 2020: intense surveillance is vital for preventing sustained transmission in new locations. Journal of clinical medicine, 9(2):498, 2020.

[26] Adam J Kucharski, Timothy W Russell, Charlie Diamond, Yang Liu, John Edmunds, Sebastian Funk, Rosalind M Eggo, Fiona Sun, Mark Jit, James D Munday, et al. Early dynamics of transmission and control of covid-19: a mathematical modelling study. The lancet infectious diseases, 2020.

[27] Andrea L Bertozzi, Elisa Franco, George Mohler, Martin B Short, and Daniel Sledge. The challenges of modeling and forecasting the spread of covid-19. arXiv preprint 2004.04741, 2020.

[28] Ronald A Fisher, A Steven Corbet, and Carrington B Williams. The relation between the number of species and the number of individuals in a random sample of an animal population. The Journal of Animal Ecology, pages 42–58, 1943.

[29] Maurice H Quenouille. A relation between the logarithmic, poisson, and negative binomial series. Biometrics, 5(2):162–164, 1949.

[30] David Applebaum. Lévy processes and stochastic calculus. Cambridge university press, 2009.

[31] M. E. J. Newman, S. H. Strogatz, and D. J. Watts. Random graphs with arbitrary degree distributions and their applications. Physical Review E, 64(2), Jul 2001.

[32] Stephen A Lauer, Kyra H Grantz, Qifang Bi, Forrest K Jones, Qulu Zheng, Hannah R Meredith, Andrew S Azman, Nicholas G Reich, and Justin Lessler. The incubation period of coronavirus disease 2019 (covid-19) from publicly reported confirmed cases: estimation and application. Annals of internal medicine, 172(9):577–582, 2020.

[33] Mingyuan Zhou and Lawrence Carin. Negative binomial process count and mixture modeling. IEEE Transactions on Pattern Analysis and Machine Intelligence, 37(2):307–320, 2013.

[34] The New York Times. https://www.nytimes.com/interactive/2020/us/coronavirus-us-cases.html, 2020.

[35] Martyn Plummer. Jags: A program for analysis of bayesian graphical models using gibbs sampling, 2003.

